# Dose-Response after Low-dose Ionizing Radiation: Evidence from Life Span Study with Data-driven Deep Neural Network Model

**DOI:** 10.1101/2024.04.09.24305578

**Authors:** Zhenqiu Liu, Igor Shuryak

## Abstract

Accurately evaluating the disease risks after low-dose ionizing radiation (IR) exposure are crucial for protecting public health, setting safety standards, and advancing research in radiation safety. However, while much is known about the disease risks of high-dose irradiation, risk estimates at low dose remains controversial. To date, five different parametric models (supra-linear, linear no threshold, threshold, quadratic, and hormesis) for low doses have been studied in the literature. Different dose-response models may lead to inconsistent or even conflicting results.

In this manuscript, we introduce a data-driven deep neural network (DNN) model designed to evaluate dose-response models at low doses using Life Span Study (LSS) data. DNNs possess the capability to approximate any continuous function with an adequate number of nodes in the hidden layers. Being data-driven, they circumvent the challenges associated with misspecification inherent in parametric models. Our simulation study highlights the effectiveness of DNNs as a valuable tool for precisely identifying dose-response models from available data. New findings from the LSS study provide robust support for a linear quadratic (LQ) dose-response model at low doses.

While the linear no threshold (LNT) model tends to overestimate disease risk at very low doses and underestimate health risk at relatively high doses, it remains a reasonable approximation for the LQ model, given the minor impact of the quadratic term at low doses. Our demonstration underscores the power of DNNs in facilitating comprehensive investigations into dose-response associations.

## Introduction

Ionizing radiation (IR) refers to radiation with sufficiently high energy to eject electrons from an atom or molecule, thereby creating ions. Both electromagnetic radiation (X- and γ-rays) and particle radiation (alpha-particles, electrons, or protons) fall under the category of ionizing radiation, as described in the WHO and BEIR VII reports [1-2]. Exposure to ionizing radiation, whether from natural or man-made sources, is inevitable in various environments. Major sources of natural radiation exposure include air, cosmic rays, food, or water [3], while exposure to artificial sources may vary within the population. Those at a higher risk of man-made radiation exposure include medical personnel, nuclear industry workers, individuals residing in air-contaminated regions, and patients undergoing medical procedures [4]. In the United States, the primary artificial sources of radiation are medical procedures like CT and X-ray scans, followed by indoor radon exposures and other anthropogenic sources [5]. According to the National Council on Radiation Protection and Measurements (NCRP), the average annual radiation dose in the U.S. currently stands at 6.3 mSv, having doubled over the past 20 years [6]. Accurately assessing the health risks associated with low-dose radiation (below 100 mGy) is crucial for establishing a scientific foundation for radiation protection and safety. Despite the well-established deleterious effects of high-dose IR, the health risks of low-dose IR remain controversial and subject to extensive debate [7, 8].

Epidemiological studies represent a widely employed approach to assess the health effects of radiation on humans, with a primary focus on comparing cancer incidences or mortalities between exposed and control groups. The Excessive Relative Risk (ERR) measures the ratio or fold change of incidence or mortality rates above baseline, while the Excess Absolute Risk (EAR) quantifies the number of cases above the control group. Despite the abundance of epidemiological evidence, seemingly straightforward interpretation, the primary uncertainties in health risk assessment arise when examining low doses below 100 mGy. Currently, the literature presents five controversial types of low dose-response models: linear-no-threshold, threshold, supra-linearity, linear quadratic, and hormesis (sub-linearity), each defended passionately within scientific discourse [7,10-14]. The Linear-No-Threshold (LNT) model, widely adopted by regulatory bodies for radiation risk assessment and health policy development, posits that every increment of radiation dose increases tumor risk for humans, with no designated ‘safe’ dose. In contrast, the threshold model contends that very small exposures to ionizing radiation are harmless. The supra-linearity model proposes that small doses of radiation are even more harmful than predicted by LNT, while the hormesis (sub-linearity) model suggests that radiation at very low doses can be beneficial. The linear quadratic model encompasses both supra-linearity and sub-linearity, featuring positive or negative coefficients in the linear and quadratic terms. Unfortunately, no consensus exists among scientific communities regarding these hypotheses.

Epidemiological studies have been cited as supporting various hypotheses, ranging from hormesis to supra-linearity at low doses. Notably, the life span studies (LSS) cohort from the Radiation Effects Research Foundation (RERF) has provided compelling evidence for the LNT risk model. However, LSS has also been implicated in supporting supra-linearity or the threshold model at low doses [14, 16]. Evidence from nuclear facility exposures at Hanford and Mayak plants has leaned towards supporting a threshold and LNT model, respectively [17]. Other studies examining medical exposures, home radon, and occupational radiation exposures have yielded inconclusive and even contradictory conclusions. The determination of the true model has significant implications for public decision-making and government regulations.

To date, epidemiological studies have predominantly relied on parametric risk models, where risk estimations are significantly influenced by model specification. The grouping of data, selection of parametric models, and choice of statistical tests, even with identical data, can markedly impact conclusions drawn about low-dose risks. In light of this, our pilot study introduces a novel approach using a data-driven deep neural network for health risk estimation.

The deep neural network (DNN) model possesses the ability to approximate any differentiable function with an adequate number of nodes in the hidden layers, as outlined in the universal approximation theory [18-20]. Characterized by its nonlinearity and independence from specific parametric settings, the DNN learns the underlying dose-response relationship solely from data. This characteristic proves invaluable in overcoming the limitations associated with parametric models when assessing tumor risk at low doses.

In our pilot study, the DNN serves as a powerful tool for exploring the dose-response relationship using life span studies (LSS) data and reconstructing the underlying dose-response model at low doses. By virtue of its flexibility and capacity to adapt to complex relationships, the DNN may contribute to determining the types of dose-response models and guiding the accurate reconstruction of parametric risk models at low doses.

## Material and Methods

### Simulation Datasets

We generated simulation data with known parameters and various dose-response models. The objective was to assess the ability of the DNN model to accurately fit and describe dose responses using simulated data derived from “true” models of different shapes.

### Real Dataset

The cancer incidence data for the life-span study (LSS) covering the 1958-1998 follow-up period was acquired from the publicly available resources at https://www.rerf.or.jp/en/ [21]. The dataset comprises a person-year summary table containing 25,570 cells, encompassing 2,764,735 person-years of follow-up for 17,448 solid tumors among 105,427 survivors. Stratification of the person-year table was conducted based on age at exposure, attained age, time since exposure, gender, city, and NIC (Not in City). For radiation dose estimation in this study, a person-year weighted, adjusted, and truncated DS02 colon dose was utilized.

### Deep Neural Network

We introduced a deep neural network approach for radiation risk prediction as illustrated in Figure 1.

**Figure 1:**
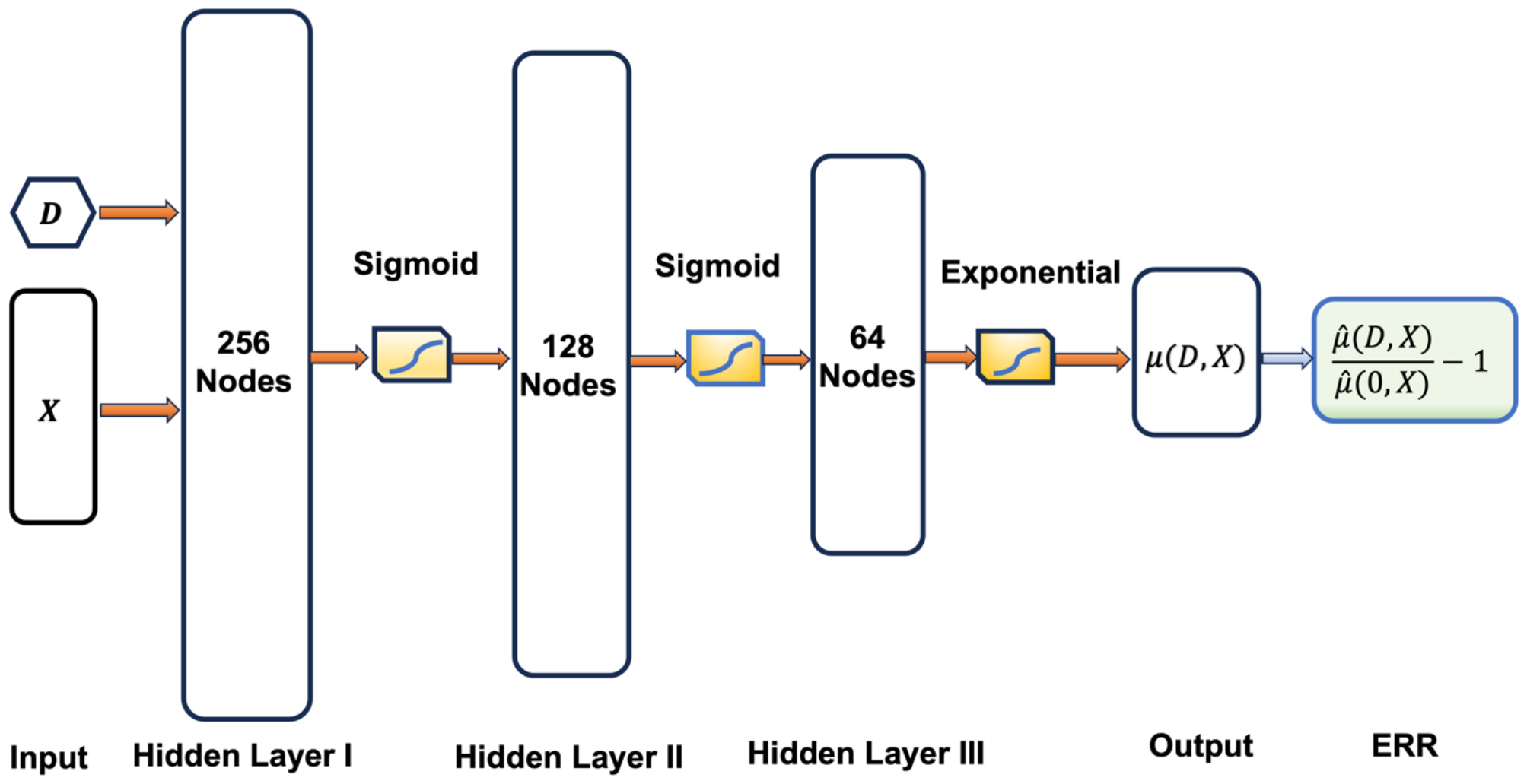
Neural network structure for radiation risk prediction, where D and X represent the radiation dose and confounding factors, respectively. ERR stands for excess relative risk. The architecture includes 3 hidden layers with node counts of 256, 128, and 64, respectively. Both sigmoid and exponential activation functions were employed. The estimation of ERR is indicated in the rightmost shaded box, determined after model training.

Given the radiation dose D, associated factors X, cancer incidence Y, and person-year Py, we build a neural network featuring 3 hidden layers with node counts of 256, 128, and 64, respectively. The selection of the number of nodes and layers is based on achieving the smallest test error. Assuming a Poisson distribution, for each cell i,

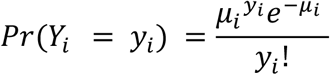

and

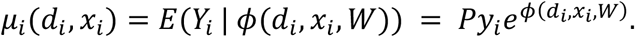

The loss function corresponds to the negative log-likelihood (NLL) of a Poisson distribution, formulated as:

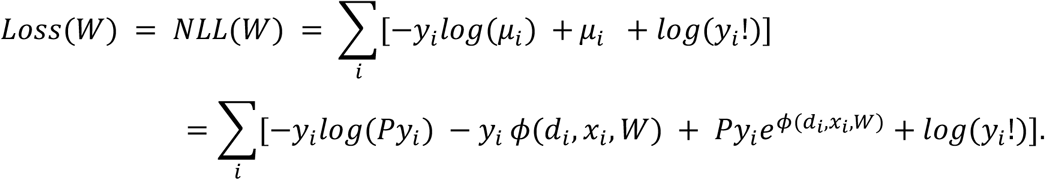

After training the model, the excess relative risk (ERR) is estimated as

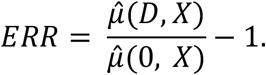

The software is implemented in Python and TensorFlow. The determination of the number of nodes and layers is achieved through cross-validation, seeking the smallest mean negative log-likelihood of the validation data. A learning rate of 0.0001 is set, and the widely used Adam optimization algorithm is employed for parameter estimation. Adam leverages adaptive learning rates, tailoring individual rates for each parameter. The actual learning rate of Adam in each iteration is bounded by the step size hyperparameter (0.0001), which is a crucial parameter to select. We recommend experimenting with step sizes of 0.001, 0.0001, and 0.00001 based on the nature of the problem at hand.

### Dose-Response Models at Low Dose

There are five potential radiation risk models following low-dose radiation exposure, including Supra-linearity, Linear No Threshold (LNT), Threshold, Linear-quadratic, and Hormesis, as illustrated in Figure 2.

**Figure 2:**
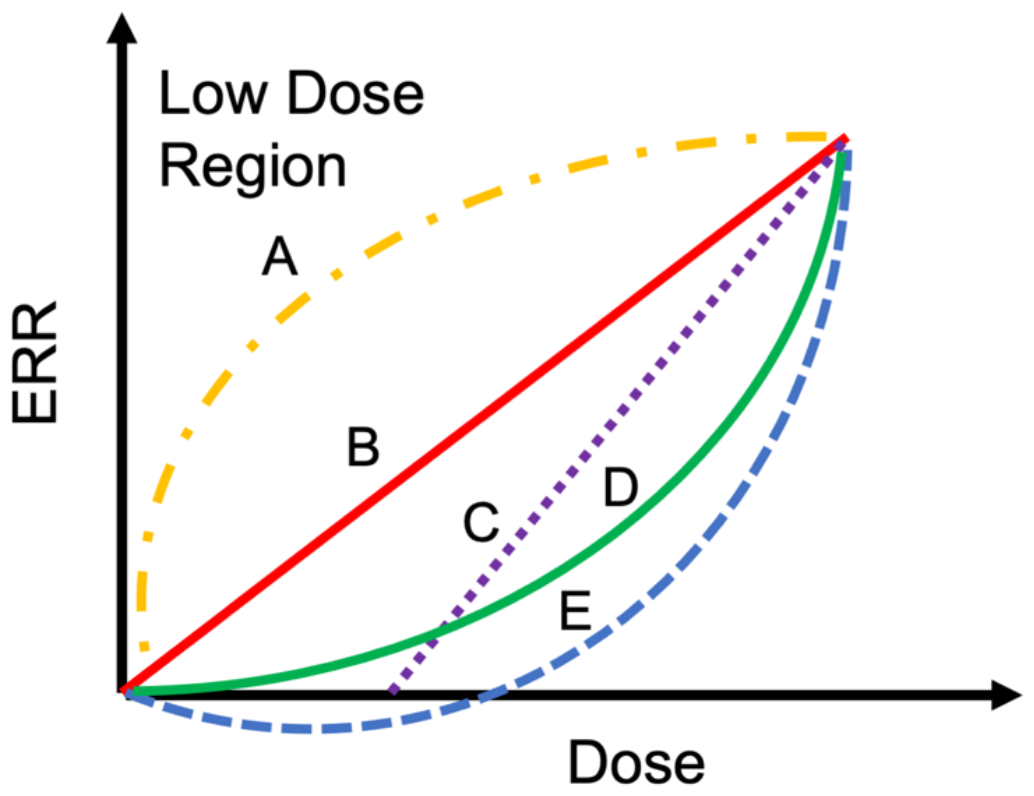
Different risk models for ERR at low dose. A: Supra-linearity, B: Linear no threshold (LNT), C: Threshold, D: Linear quadratic (LQ), E: Hormesis.

Note that at low doses, the supra-linearity model exhibits a positive ERR with a decreasing gradient (slope) concerning dose. In contrast, the Linear no-threshold model demonstrates a positive ERR with a constant gradient (partial derivative k) in relation to dose. The Threshold model indicates zero (or near-zero) ERR and zero gradients at low doses. The Linear quadratic model features a positive ERR with a linear gradient. The hormesis model, on the other hand, displays both a negative ERR and an increasing gradient at low doses. Consequently, we establish the following straightforward rules at very low doses:

A. Supra-linearity, if

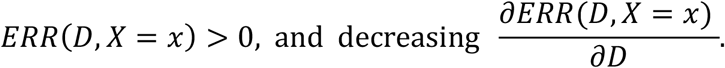
B. Linear no-threshold (LNT), if

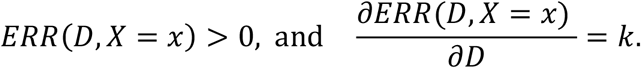
C. Threshold, if

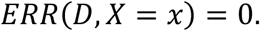
D. Quadratic, if

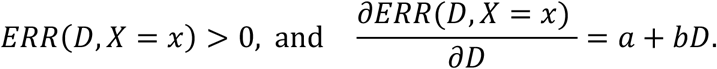
E. Hormesis, if

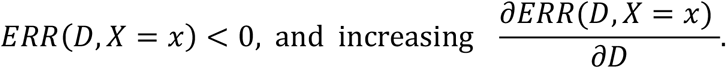

### Numerical Gradient Computation

Excess Relative Risks (ERRs) at a specific dose can be directly computed from the DNN model. However, the first-order derivatives need to be estimated numerically [22]. In this study, we employed the following two numerical differentiation methods.

i. Central Difference (2^nd^ order):

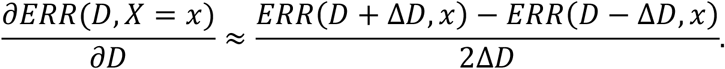
ii. Central Difference (4^th^ order):

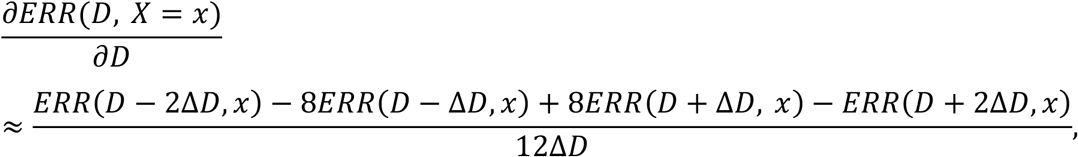

where Δ*D* is the step size. There are more accurate approximations in the Python package [22], but we did not detect any significant differences with different numerical methods.

### Simulations

We conducted simulations for low-dose IR exposure using data generated from known model structures and various dose-response functions, as outlined below:

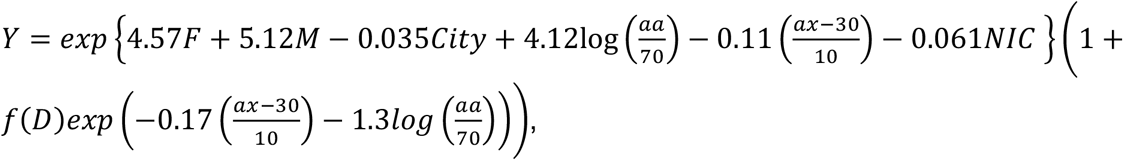

where F: Female, M: Male, City: Hiroshima (Yes/No), aa: attained age, ax: age at exposure, NIC: not in the city (Yes/No), and Y: Incidence rate. The coefficients in the simulation model were initially derived from a linear dose-response model using person-year summary data [1]. Attained age and age at exposure are randomly generated from the person-year table within the ranges of 13-101 and 0-80, respectively. The computational results, encompassing various dose-response functions (f(D)s), attained ages, and age at exposures, are presented in Figures 3 and Supplementary Figure 1, respectively.

**Figure 3:**
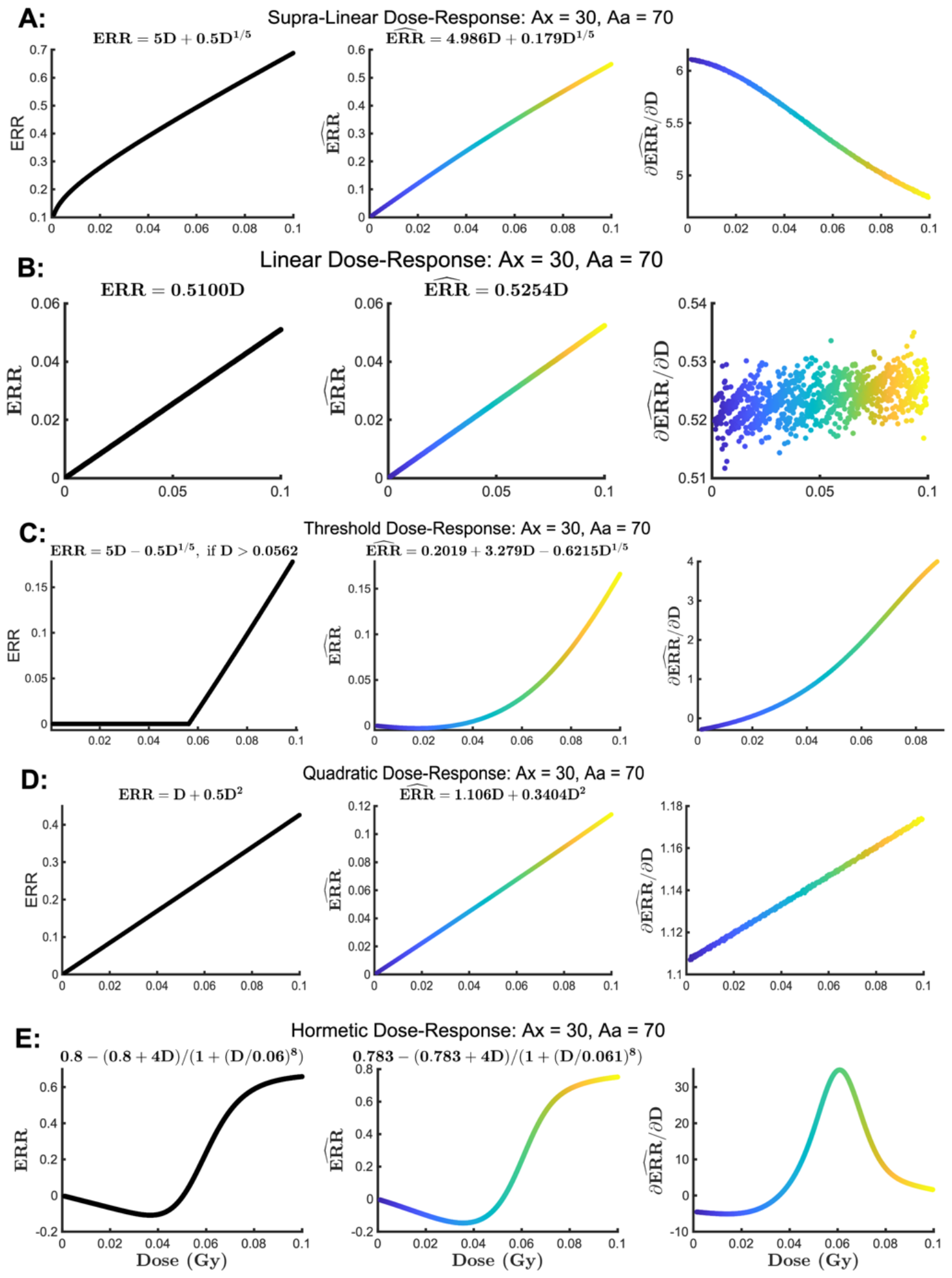
Plots of true ERRs and DNN-estimated ERRs, along with their first-order derivatives, for various dose-response models, with the age at exposure and attained age set at 30 and 70, respectively. Left panels: Plots of true ERRs with different dose-response functions (f(D)s); Middle panels: Plots of the estimated ERRs using DNN; Right panels: Plots of the first-order derivatives obtained from DNN. Figures 3 A-E represent the dose-response models of supra-linear, Linear No-Threshold (LNT), threshold, quadratic, and hormesis, respectively.

As depicted in Figure 3, the dose-response functions (f(D)s) for supra-linear, Linear No-Threshold (LNT), threshold, linear quadratic, and hormesis models are given by

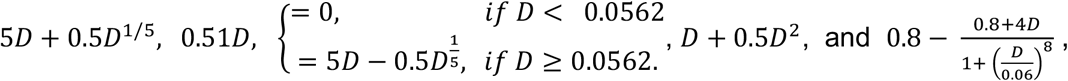

respectively. Note that we deliberately assigned much smaller nonlinear coefficients in each model to assess the DNN’s capability to detect nonlinear effects and accurately capture true dose-response relations. In our simulation model, the true Excess Relative Risk (ERR) is represented as f(D) with an attained age of 70 and an age at exposure of 30, where the age-modified term is 1. Particularly, in Figure 3A, moving from left to right, the plots represent supra-linear patterns for true ERR, estimated ERR, and estimated ^∂*ERR*^/_∂*D*_. We subsequently refitted a nonlinear regression using the estimated ERR and known dose, as shown in the middle panel. The resulting estimated ERR is 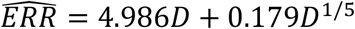 (adjusted *R*^2^ = 0.9997). In comparison to the true model (*ERR* = 5*D* + 0.5*D*^1/5^), the estimated coefficient of the linear term (4.986) closely approximates its true value of 5, while the estimated coefficient of the nonlinear term (0.179) is smaller than the true value of 0.5. Nonetheless, the DNN accurately identifies the supra-linear model type, with an estimated 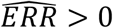 and decreasing 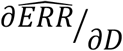 at low doses, as illustrated in the middle and right panels of Figure 3A. Similarly, in Figure 3B, we obtained the estimated ERR as 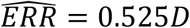 (adjusted *R*^2^ = 1) after re-fitting the linear model with the estimated ERR from DNN (middle panel), which closely aligns with the true model of 0.51D. The 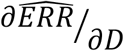 appears akin to white noise unrelated to the dose, with a mean around 0.525 (right panel). Despite the challenge of obtaining a perfectly horizontal line for 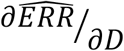 due to roundoff errors and approximations, the right panel of Figure 3B indicates that 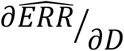 is a constant across different doses. Once again, DNN accurately identified the linear model type with a straight line in 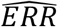 and a constant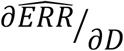 . Furthermore, in Figure 3D, DNN accurately identifies the quadratic model type with a positive ERR and a linear 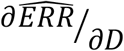 (right panel), despite the estimated model coefficients being slightly different as 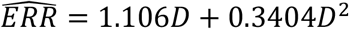 (adjusted *R*^2^ = 1) compared to the true model *D* + 0.5*D*^2^. Moreover, in Figure 3E, DNN appropriately recognizes the hormesis model type with a negative 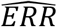and an increasing 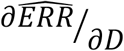 at very low doses. The estimated model from DNN, 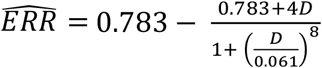 (adjusted *R*^2^ = 0.9987), closely resembles the true model 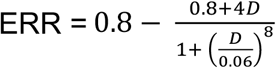. Finally, detecting a threshold model is somewhat challenging due to its lack of full differentiability. As illustrated in the middle panel of Figure 3C, the most suitable continuous function for the estimated ERR from DNN is given by 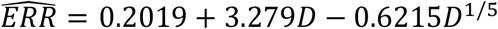(adjusted *R*^2^ = 0.9382), which partially captures the nonzero part of the true threshold model:

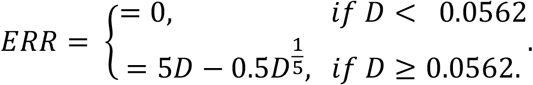

Nevertheless, DNN still presents compelling evidence for a threshold model, as 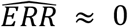 at very low doses (middle panel of Figure 3C). Similar observations with different ages at exposure are noted in Supplementary Figure 1. Overall, DNN proves to be more efficient in distinguishing differentiable dose-response models (supra-linear, linear, linear-quadratic, and hormesis), but it exhibits some inefficiency in detecting a threshold model due to the lack of full differentiability in the model. In general, it remains a valuable tool for uncovering the underlying type of dose-response models.

### LSS Cohort Data

We constructed a feed-forward DNN model with one input, one output, and three hidden layers, as depicted in Figure 1. The first, second, and third hidden layers consist of 256, 128, and 64 nodes, respectively. The selection of the number of nodes and hidden layers is based on minimizing the average negative log-likelihood. In addition to radiation dose, six other covariates—namely, attained age, age at exposure, time since exposure, sex, city, and NIC—are incorporated into the model. The dataset was randomly split into 80% training and 20% validation data. Prior to input into the DNN, all variables were normalized to have zero means and standard deviations of 1. The corresponding output represents solid tumor incidence.

After training the model, we generated test data and estimated the 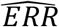 and 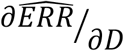 at low doses (< 100 mGy). The computational results are presented in Figure 4.

**Figure 4:**
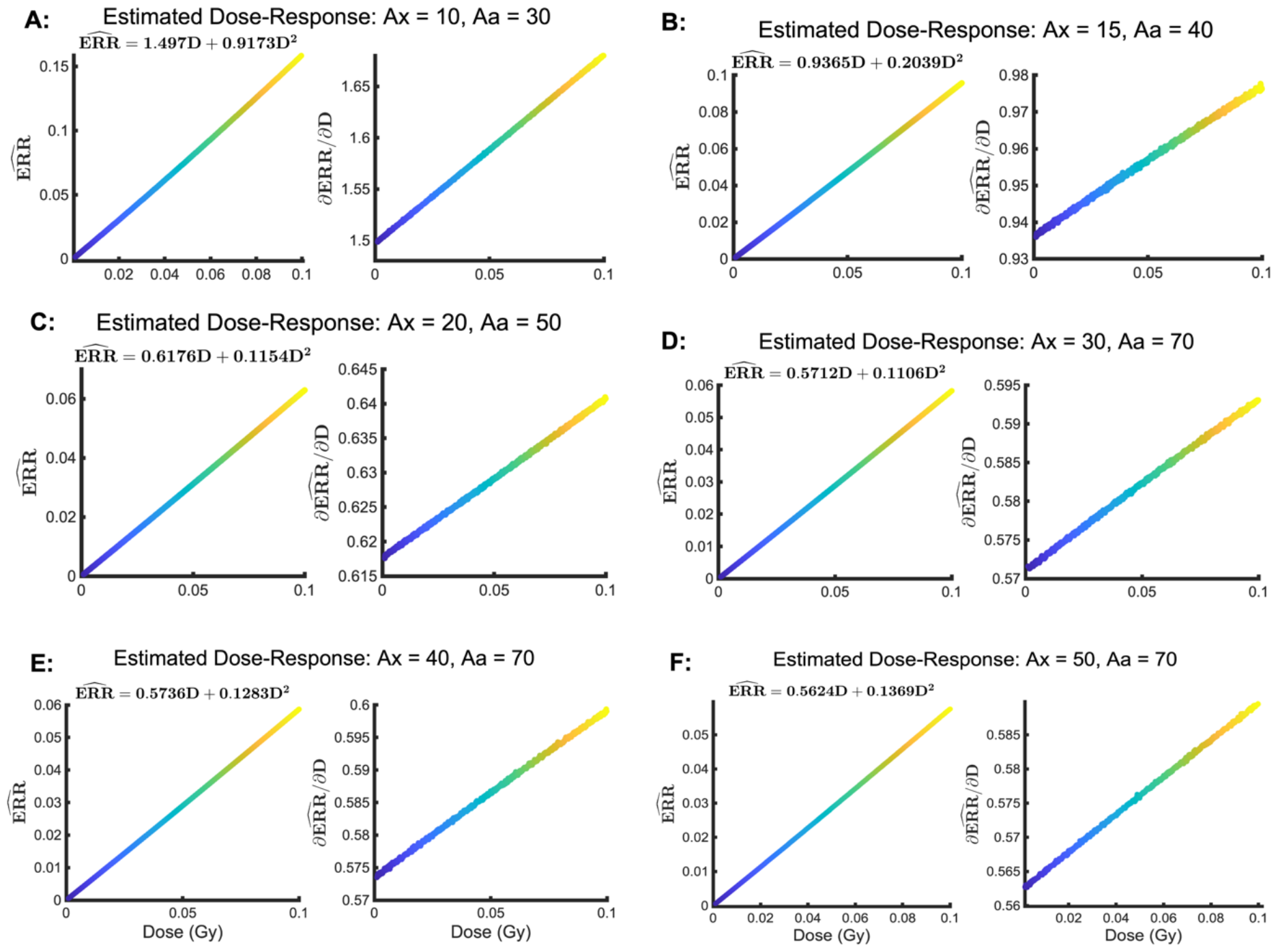
Estimated Excess Relative Risk (ERR) and its first-order partial derivative for different ages at exposure (Ax) and attained ages (Aa) for a woman residing in Nagasaki. A: Ax = 10 and Aa = 30; B: Ax = 15 and Aa = 40; C: Ax = 20 and Aa = 50; D: Ax = 30 and Aa = 70; E: Ax = 40 and Aa = 70; F: Ax = 50 and Aa = 70.

Figures 4A-F consistently reveal a quadratic dose-response relationship across different ages at exposure and attained ages in the LSS cohort. For example, in Figure 4A, a linear 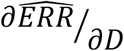 is detected (left panel), and fitting a quadratic model (*a* + *bD* + *cD*^2^) with the estimated ERR yields an estimated dose-response model of 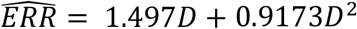 with an adjusted *R*^2^ = 1, indicating nearly perfect fitting. Similar results are observed with other ages at exposure and attained ages (Figures 4B-4F), suggesting a Linear quadratic (LQ) model with the LSS cohort data. While the coefficients of both linear and quadratic terms diminish with increased age at exposure and/or an older attained age, the LQ model remains the most fitting dose-response model for the data, consistently achieving a perfectly adjusted *R*^2^ *of* 1 for different age groups. Despite potential bias in the estimated coefficients relative to the true model, as observed in the simulation, DNN accurately identified the type of quadratic dose-response model. Additionally, Supplementary Figure 2 indicates that a log-linear dose-response model (*ERR* = *ae*^*bD*^) is not a valid option for the LSS cohort data, as the log(*ERR*) exhibits a supra-linear trend with different ages.

## Discussion and Conclusions

Accurately assessing the health risk of low-dose radiation is crucial for establishing a scientific foundation for radiation protection and safety. Parametric risk models, extensively studied and employed by academics and government agencies, offer simplicity and ease of interpretation. However, designing these models requires substantial domain expertise, and mis-specifying them may result in bias and inaccurate risk estimation. In contrast, data-driving DNNs prove highly efficient and robust, capable of approximating any underlying continuous functions and achieving state-of-the-art performance without the constraints of predefined function forms. Nevertheless, the inherent challenge with DNNs lies in their black-box nature, making their internal decision-making processes opaque and challenging to interpret. This lack of transparency raises significant concerns, particularly in critical applications such as healthcare, where understanding the reasoning behind a decision is essential.

To bridge the gap between the remarkable performance of DNNs and the comprehensibility of parametric models, it is valuable and of strong interest to identify the type of a parametric dose-response model from data with the guidance of DNNs. This pilot study represents our initial effort in this direction. DNNs, free from the problem of model misspecification, can be employed to discern different dose-response models at low doses and guide the construction of a correct parametric risk model.

Our simulations, employing known dose-response functions and model structures, highlight that DNN offers a valuable approach for discerning types of dose-response models. In our analysis of real data from the LSS cohort, we identify a LQ dose-response model, contrasting with a linear no-threshold (LNT) model. While the LNT model may overestimate the risk at very low doses, it tends to underestimate the risk at relatively high doses. Nonetheless, the LNT model serves as a reasonable approximation for LQ, given the minor contribution of the quadratic term at very low doses.

DNN proves effective in identifying and analyzing complex relationships between doses and responses, playing a pivotal role in providing valuable insights into various types of dose-response models. Additionally, it uncovers subtle nuances in dose-response relationships that might be overlooked by traditional statistical approaches. Consequently, DNN holds significant potential to advance our understanding of dose-response modeling, enabling more accurate predictions and enhancing decision-making in the realm of radiation protection.

## Data Availability

All data produced in the present study are available upon reasonable request to the authors

## ACKNOWLEDGMENTS

The Radiation Effects Research Foundation (RERF), Hiroshima and Nagasaki, Japan is a private, nonprofit foundation funded by the Japanese Ministry of Health, Labour and Welfare (MHLW) and the U.S. Department of Energy (DOE). The research was also funded in part through the DOE award DE-HS0000031 to the National Academy of Sciences. The views of the authors do not necessarily reflect those of the two governments.

---

**Supplementary Figure 1:**
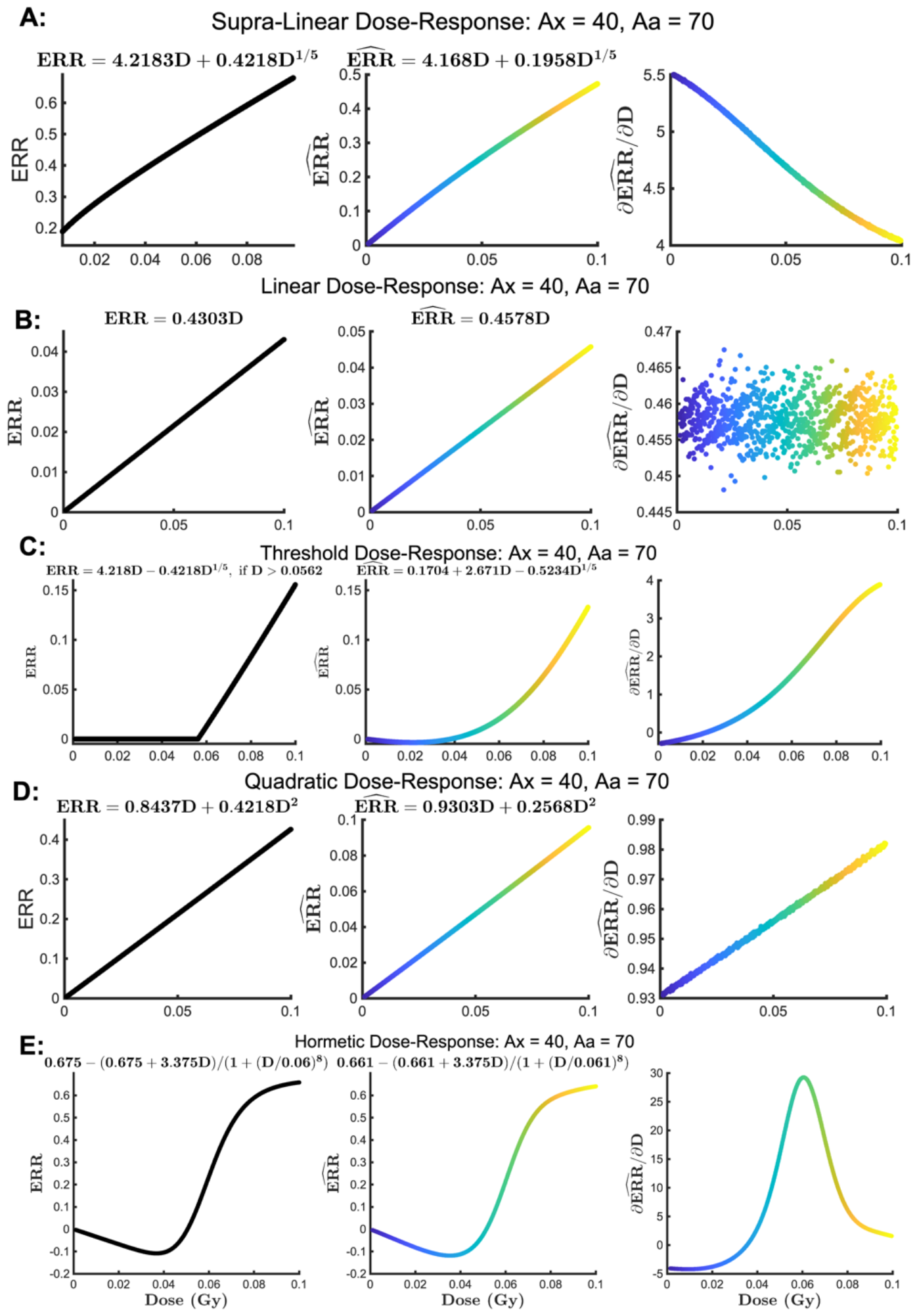
Plots of True Excess Relative Risks (ERRs) and Deep Neural Network (DNN) Estimated ERRs, Along with their First-Order Derivatives, for Various Dose-Response Models at Ages 40 and 70. Left Panels: Plots of the True ERRs with Different Functional Forms (f(D)). Middle Panels: Plots of the Estimated ERRs using DNN. Right Panels: Plots of the First-Order Derivatives from DNN. Supplementary Figures 1A-E depict the dose-response models of supra-linear, Linear No-Threshold (LNT), threshold, linear quadratic, and hormesis, respectively, with consideration of the age at exposure and attained ages of 40 and 70.

**Supplementary Figure 2:**
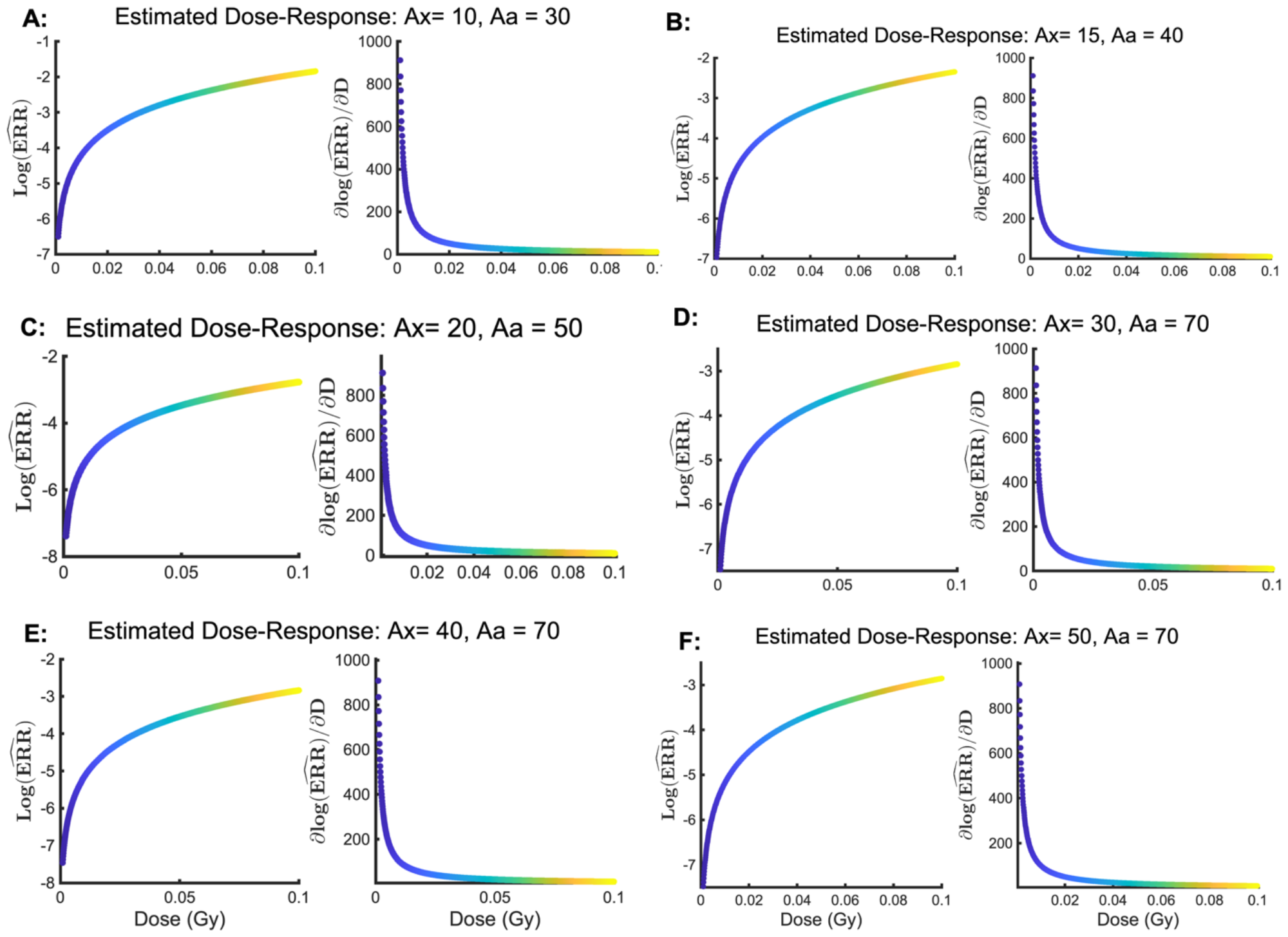
Estimated Logarithm of Excess Relative Risk (log(ERR)) and its First-Order Partial Derivative for Various Ages at Exposure (Ax) and Attained Ages (Aa) in a Woman Residing in Nagasaki. Panel A: Ax=10 and Aa=30; Panel B: Ax=15 and Aa=40; Panel C: Ax=20 and Aa=50; Panel D: Ax=30 and Aa=70; Panel E: Ax=40 and Aa=70; Panel F: Ax=50 and Aa=70.

